# First detection of Monkeypox virus genome in sewersheds in France

**DOI:** 10.1101/2022.08.18.22278938

**Authors:** Sebastien Wurtzer, Morgane Levert, Eloise Dhenain, Mickael Boni, Jean Nicolas Tournier, Nicolas Londinsky, Agnès Lefranc, Obepine SIG, Olivier Ferraris, Laurent Moulin

## Abstract

A monkeypox virus outbreak is currently spreading in multiple non-endemic countries since May 2022. The atypical clinical profile of patients has led to a very likely underestimation of the number of cases at the beginning of epidemic. The detection and quantification of the Monkeypox virus genome in sewersheds in Paris (France) correlated temporally with the identification of the first case of infection and the spread of the disease within the population connected to the sewage system.

## Main text

Since May 2022, a new zoonotic infectious disease, Monkeypox, has gained attention of health authorities after starting to circulate in wealthy countries usually spared. Monkeypox is caused by Monkeypox virus (MPXV), a member of the *Orthopoxvirus* genus of the *Poxviridae* family, and results in pox-like skin lesions, making the diagnosis difficult with smallpox and chickenpox virus infection. This infection is becoming endemic in a dozen countries in West and Central Africa ^1^. Human infections with the clade 1 (former Congo Basin clade) appeared to cause more severe disease compared to the clade 2 and 3 ^1–3^. Genomic sequencing of MPXV implicated in the 2022 outbreak have determined its relationship to the clade 3 (former West African clade) ^3–5^. This Monkeypox outbreak was reported on May 7th, 2022 in the United Kingdom ^6^. Since May 13th, 2022, Monkeypox cases have been reported to WHO in 12 member states for which Monkeypox was not endemic ^7^. Epidemiological investigations are underway to understand the routes of transmission.

It is likely that this number of cases is greatly underestimated in countries where circulation is endemic ^8^ and, as surveillance is intensified in non-endemic areas, new cases may be identified. In France, Monkeypox disease are subject to ongoing surveillance through mandatory reporting. On July 12th, 2022, 912 cases of Monkeypox were officially reported in France, 569 of which were in the Greater Paris region ^9^.

MPXV infection starts with very general symptoms followed by vesicular eruptions appear about 2 days after the onset of the infection but the incubation period of the disease can be from 5 to 21 days, making the identification of contamination complex ^10^. Little information has been reported on the virus excretion kinetics during the infection in various biological fluids that have to be analyzed. However it has been established that virus genome could be detected in skin lesions, feces, saliva, urine and semen for prolonged period (16 days since symptom onset) ^11^.

In the current context, the key objectives of surveillance and case investigation are to identify isolated cases, potential clusters and the infection origin as soon as possible in order to provide clinical care and isolate cases to prevent transmission. Containment of the virus circulation is therefore mainly based on early infection diagnosis, isolation of patient, and vaccination of the population at risk. However, this approach requires medical consultation and adherence to isolation measures of patients. The biological diagnosis carried out on people who underwent other sexually transmitted infections showed the possibility of asymptomatic carriers or patients presenting an atypical clinical presentation ^12^, capable of transmitting the virus, thus suggesting an underestimation of the viral circulation through the symptomatic case surveillance ^8,13,14^.

Since 2020, interest in wastewater-based epidemiology (WBE) has considerably increased with the detection and quantification of the SARS-CoV-2 genome in raw wastewater ^15^. This approach is made possible because SARS-CoV-2 is shed in the stool of infected persons ^16^, even if they are pre-, pauci- or asymptomatic. Numerous studies have demonstrated the correlation between the incidence of the disease, the positivity rate of tests and the concentration of viral genomes in wastewater ^17–19^.

The objectives of this work were to demonstrate the presence of MPXV genome in the sewersheds in the city of Paris (France), and to date the virus emergence. Monkeypox WBE could be a supplementary tool for the health authorities to better understand the viral circulation within the population.

For more than 2 years, 16 sewersheds located in the city of Paris have been weekly sampled during for 24h. First detection of MPXV genome occurred in wastewater on May 23rd, 2022 in 3 different sewersheds in Paris (figure 1). The first French human case was officially reported by May 19th, 2022 in Paris and 3 human cases were reported on May 23rd, 2022. Based on compliance with isolation measures of the first Monkeypox-diagnosed patients, genome detection of in sewersheds covering other areas through the following weeks could suggest that other cases might have existed and not been diagnosed yet when first human cases were identified. Out of the 16 sewersheds under investigation, the fraction of positive ones for MPXV genome increased from 3/16 (May 23rd) to 9/16 on July 11th, 2022 indicating the virus spread in the population connected to the sewage network (figure 2A). The results were in accordance with the continuous increase in new human cases officially reported each week ^20^.

**Figure 1.**
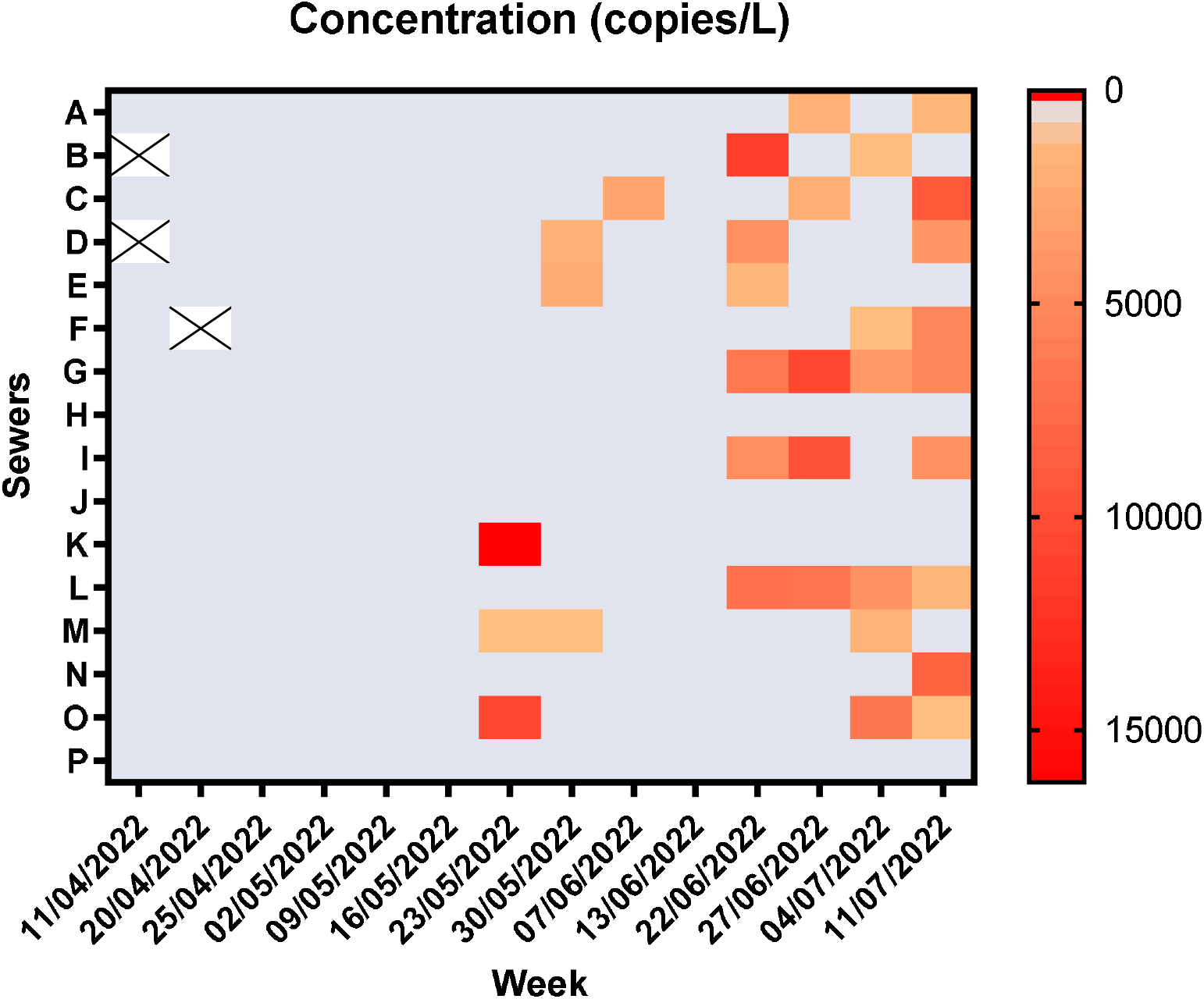
Heatmap of the MPXV genome concentration in wastewater collected weekly in 16 sewersheds in the city of Paris, France. Grey for non-detected genome and X when the sample was not available.

**Figure 2.**
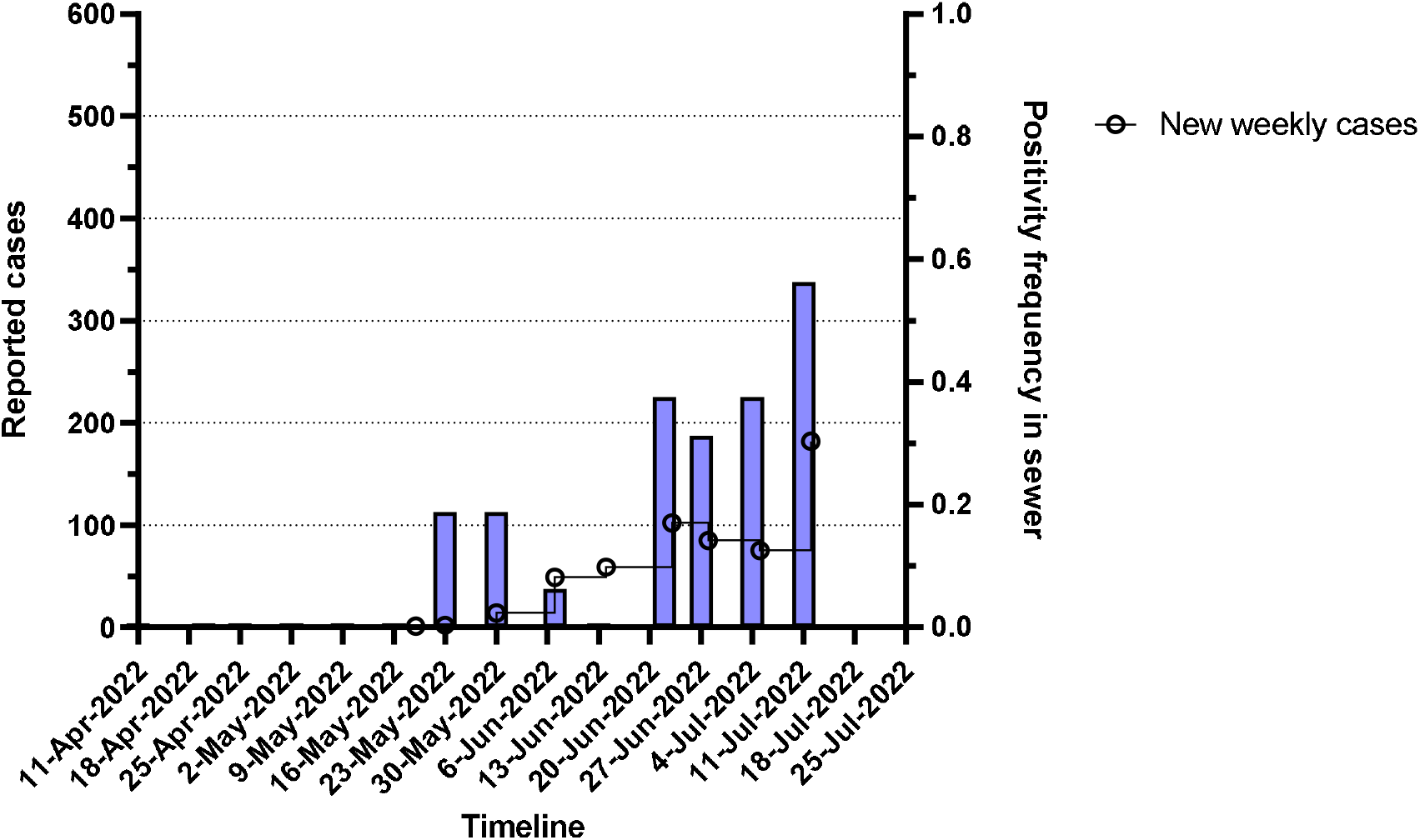
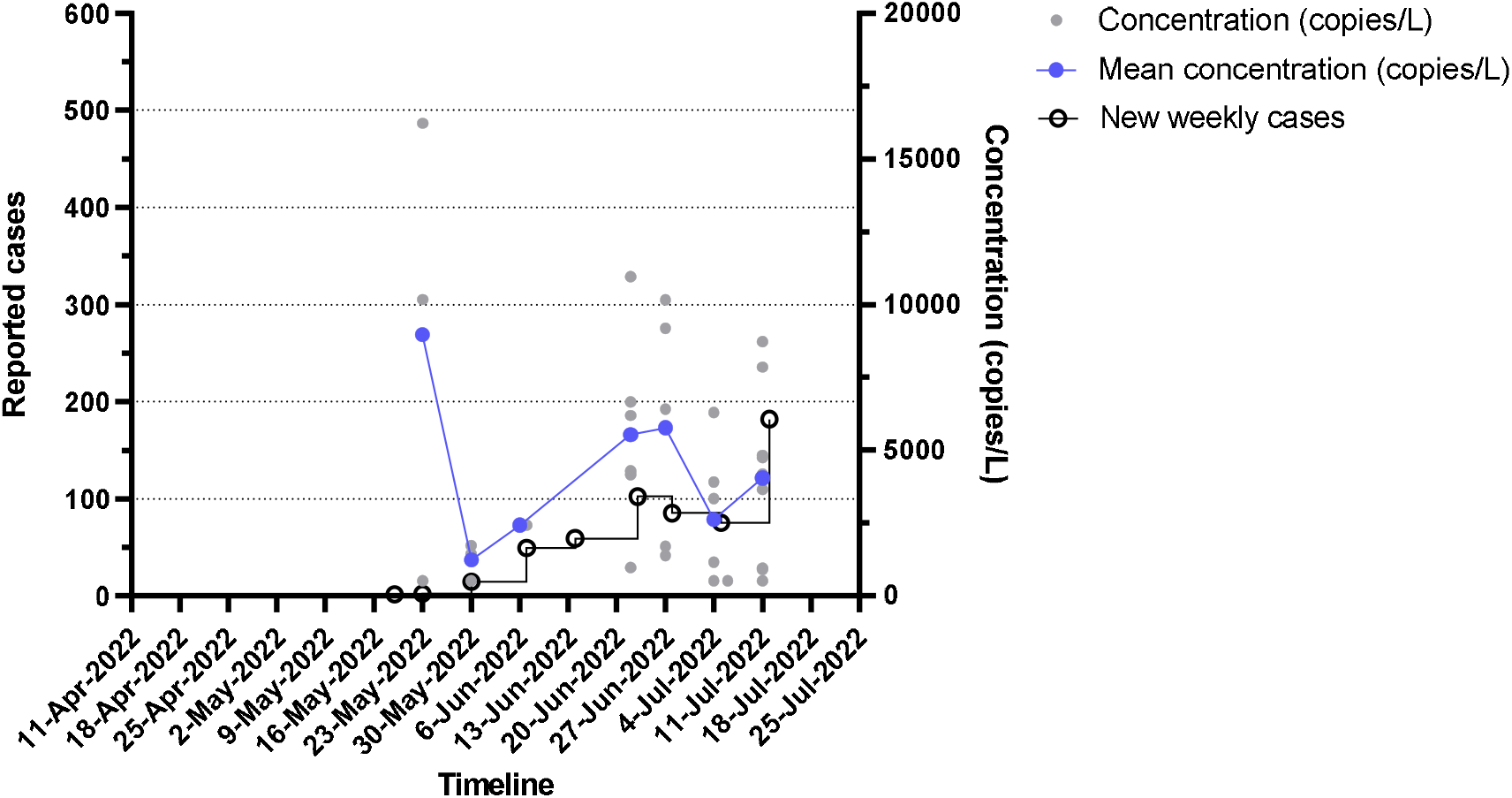
A: Frequency of positivity in sewershed samples in Paris from April 11th to July 11th, 2022. New weekly human cases have been reported in open circles. B: Quantification of the MPXV genome concentration in sewer sampled in Paris from April 11th to July 11th, 2022. Viral concentration of positive samples in grey dot point, average viral concentration in purple dot point and new weekly human cases in open circles.

Estimated MPXV genome concentrations were globally between 1,000 and 10,000 copies/L for less than 1,000 reported infected patients (figure 2B). Excepted the first positive samples, average MPXV genome concentration increased concomitantly with the number of new weekly human cases. Viral genomes found in raw wastewater may originate mainly from viruses excreted in body fluids but also from viruses contained in skin lesions released during hand and body washing ^11^. With all the usual precautions regarding the comparison of Ct values, the genome quantities detected in the biological samples likely to contaminate the wastewater were of the same order of magnitude in COVID-19 and Monkeypox patients ^11,21,22^. Comparisons of SARS-CoV-2 and MPXV concentrations in wastewater were difficult to understand because of the high underestimation of COVID-19 cases at the beginning of the pandemic. In addition, contrary to biological fluids, viral genomes in scabs may be more protected from degradation but may also be responsible for a less homogeneous viral concentration in wastewater samples leading to a « nugget » effect such the one observed in sewersheds sampled by May 23rd, 2022.

MPXV strains from the two worldwide outbreaks (2017-2022) belonged to MPXV clade 3 ^23^. The low severity during the 2022 MPXV outbreak and asymptomatic carriers may have led to erroneous clinical diagnoses. Then, a silent circulation of the MPXV in naïve populations no longer immune to smallpox was possible. To address this question, we conducted a retrospective analysis of randomly selected samples (n=39) from August 2021, September 2021 and March 2022. MPXV viral genome was not detected in any of them suggesting a date of the emerging event in May 2022. We could not exclude the possibility of genetic material loss resulting from freezing/thawing of extracted nucleic acids in the case of retrospective analyses. However recent phylogenetic analysis showed that MPXV genomes from the 2022 outbreak came from a new lineage called B.1 ^24^, confirming this hypothesis.

This lack of MPXV genome detection before the identification of the first human cases would also strongly suggest the human origin of the viral genomes detected in the Parisian sewersheds and the absence of a pre-existing animal reservoir for MPXV. However, the risk of contamination of peri-domestic fauna living in sewersheds have to be investigated in correlation with the study of persistence of MPXV in wastewater ^25,26^.

To our knowledge, this is the first study reporting the detection and quantification of MPXV genomes in sewersheds. This approach has allowed to date the emergence concomitantly with the first human case identified and to observe the spread of the 2022 Monkeypox epidemic in Paris (France) by wastewater monitoring (n=264 samples) over a 10-month retrospective period, highlighting once again the importance of WBE to establish an early warning system for epidemic emergence. The interest of retrospective analyses to understand the emergence of epidemics pointed out the fundamental need for wastewater sample banks. For the time being, the concentration of genomes in wastewater appeared to be relatively low. Routine monitoring will be helpful to establish the viral spread in the population as well as shedding kinetics in patients. Considering the mode of transmission and the relatively low number of human cases, such results might be more difficult to implement than for SARS-CoV-2.

## Material & methods

### Sample collection

Sixteen sewers in the city of Paris (France) were weekly sampled since May 2020 for initially SARS-CoV-2 monitoring. Twenty-four-hours composite samples (according to NF T 90-90-523-2) were taken by automated samplers. Sampling was proportional to the flow rate, it started at 7:00 AM and finished at J+1, 7:00 AM. A minimum of 144 sub-samples per day were taken during dry weather periods. Samples were taken by suction and collected in a refrigerated polyethylene tank at 5°C (+or- 3°C). The final collected volume was between 8.7 and 14L. Then samples were carefully homogenized, distributed in a 100mL polyethylene bottle, transported to the laboratory at 4°C and processed in less than 24 hours after sampling. A total of 264 samples from sewage network of the city of Paris were processed for MPXV genome detection.

### Concentration method

All samples were processed as previously described ^18^. Briefly, samples were homogenized, then 11 ml were centrifugated at 200,000 x g for 1 hour at 4°C using a XPN80 Coulter Beckman ultracentrifuge using a swing rotor (SW41Ti). Pellets were resuspended in 200 μL of Dulbecco’s Phosphate-buffered saline (DPBS) 1x (reference 14190144, ThermoFisher Scientific) and pretreated for dissociating viruses and organic matter that was then removed from supernatant for improving RNA extraction efficiency, according to the manufacturer’s recommendations. Supernatant was then lysed, and total nucleic acids were purified using PowerFecal Pro kit (QIAGEN) on a QIAsymphony automated extractor (QIAGEN) and eluted in 50 µL of elution buffer according to manufacturer’s protocol. Even if recovery rate for MPXV could not be evaluated experimentally, this protocol performed well on different types of enveloped and naked viruses ^27^. All nucleic acids were finally purified using OneStep PCR inhibitor removal kit (Zymoresearch) according the manufacturer’s instructions and then directly used or stored at −80°C before use. The recovery rate of methods was estimated using bovine coronavirus spiked (mean recovery rate of 75%, ranging between 65% and 90%, Coefficient of Variation (CV%) of 12%) and the repeatability of the measurement was also evaluated on endogenous Pepper Mild Mottle Virus genome (CV% of 15%).

### Detection and Molecular quantification

The genome of the MPXV was detected by qPCR using the MPXV TaqMan assay (#Vi07922155_s1, ThermoFisher scientific) targeting the gene J1L. Amplification was done using Fast virus 1-step MasterMix (ThermoFisher scientific) according to the manufacturer’s instructions on Viaa7 real time thermocycler (ThermoFisher scientific). Briefly, cycling was performed as follow: polymerase activation step at 95°C for 20 sec, then amplification was done by 45 cycles of incubation at 95°C for 5 sec and 58°C for 40 sec. No template controls were included in each experiment to ensure no contamination and to set up the positivity threshold.

Some positive samples have also been amplified by digital PCR using the same MPXV TaqMan assay and QIAcuity Probe PCR Kit (QIAGEN) according to the manufacturer’s recommendations. Briefly, cycling was performed as follow: polymerase activation step at 95°C for 2 min, then amplification was done by 45 cycles of incubation at 95°C for 5 sec and 58°C for 40 sec.

Concentrations obtained by dPCR and Ct resulting from qPCR assay have been plotted to establish a standard curve allowing the quantification of all samples positive in qPCR and to determine PCR efficacy.

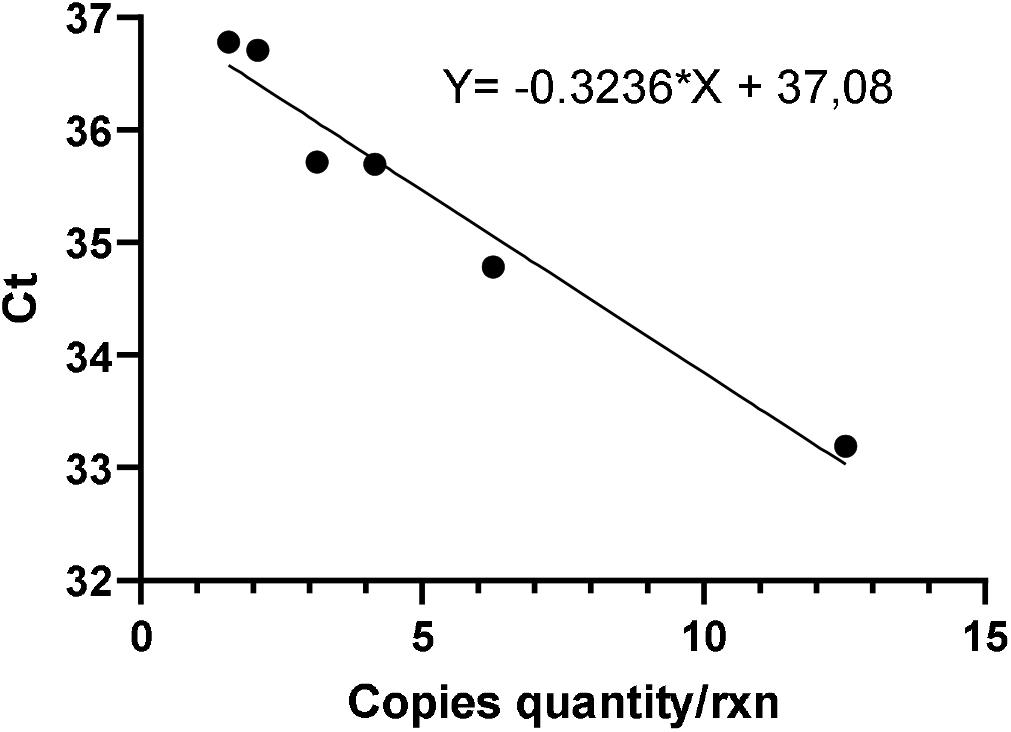

### Graphical representation

All graphics were done using GraphPad Prism software v9.0.1.

## Data Availability

All data produced in the present study are available upon reasonable request to the authors

## Author’s contribution

SW, LM, OF, MB, AL set up analytical protocols

ED, ML, SW realized the analyses

SW, ML, LM, MB interpreted the results

SW wrote the first draft

ML, ED, LM, MB, OF, JNT, AL edited the manuscript

Obepine SIG reviewed the submitted manuscript

## Data availability statement

All data produced in the present study are available upon reasonable request to the authors.

## Acknowledgments

OBEPINE scientific interest group is composed by Boni M. (Institut de recherche biomedicale des armées), Wurtzer S., Moulin L. (Eau de Paris), Mouchel JM., Maday Y., Marechal V. (Sorbonne universite), Le Guyader S. (IFREMER), Bertrand I., Gantzer C. (Universite de Lorraine)

